# Community prevalence of SARS-CoV-2 in England: Results from the ONS Coronavirus Infection Survey Pilot

**DOI:** 10.1101/2020.07.06.20147348

**Authors:** Koen B Pouwels, Thomas House, Julie V Robotham, Paul J Birrell, Andrew Gelman, Nikola Bowers, Ian Boreham, Heledd Thomas, James Lewis, Iain Bell, John I Bell, John N Newton, Jeremy Farrar, Ian Diamond, Pete Benton, Ann Sarah Walker, the COVID-19 Infection Survey team

**Author notes:** contribution considered equal (Koen B Pouwels, Thomas House). Correspondence to (or @kb_pouwels on Twitter). See Acknowledgements for the Coronavirus Infection Survey team.

## Abstract

**Objective:** To estimate the percentage of individuals infected with severe acute respiratory syndrome coronavirus-2 (SARS-CoV-2) over time in the community in England and to quantify risk factors.

**Design:** Repeated cross-sectional surveys of population-representative households with longitudinal follow-up if consent given.

**Setting:** England

**Participants:** 34,992 Individuals aged 2 years and over from 16,722 private residential households. Data were collected in a pilot phase of the survey between 26 April and 28 June 2020.

**Main outcome measures:** Percentage of individuals in the community testing positive for SARS-CoV-2 RNA using throat and nose swabs. Individuals were asked about any symptoms and potential risk factors.

**Results:** The percentage of people in private-residential households testing positive for SARS-CoV-2 reduced from 0.32% (95% credible interval (CrI) 0.19% to 0.52%) on 26 April to 0.08% (95% CrI 0.05% to 0.12%) on 28 June, although the prevalence stabilised near the end of the pilot. Factors associated with an increased risk of testing positive included having a job with direct patient contact (relative exposure (RE) 4.06, 95% CrI 2.42 to 6.77)), working outside the home (RE 2.49, 95% CrI 1.39 to 4.45), and having had contact with a hospital (RE 2.20, 95% CrI 1.09 to 4.16 for having been to a hospital individually and RE 1.95, 95% CrI 0.81 to 4.09 for a household member having been to a hospital). In 133 visits where individuals tested positive, 82 (61%, 95% CrI 53% to 69%) reported no symptoms, stably over time.

**Conclusion:** The percentage of SARS-CoV-2 positive individuals declined between 26 April and 28 June 2020. Positive tests commonly occurred without symptoms being reported. Working outside your home was an important risk factor, indicating that continued monitoring for SARS-CoV-2 in the community will be essential for early detection of increases in infections following return to work and other relaxations of control measures.

**What is already known on this topic:**

- Unprecedented control measures, such as national lockdowns, have been widely implemented to contain the spread of SARS-CoV-2.
- Previous mass surveillance has been based on data sources such as hospital admission, deaths or self-reported symptoms that do not measure community prevalence of virus directly.
- Decisions regarding the continued need for social distancing measures in the overall population, specific subgroups and geographic areas heavily rely on accurate and up-to-date information about the number of people and risk factors for testing positive.

**What this study adds:**

- The percentage of individuals from the general community in England testing positive for SARS-CoV-2 clearly declined between 26 April and 28 June 2020 from around one in three 300 to around one in a thousand.
- Risk factors for testing positive included having a job with direct patient contact, having had (indirect) contact with a hospital in the past 2 weeks, and working outside your home.
- Positive tests commonly occurred without symptoms being reported and the percentage of individuals with a positive test that reported no symptoms was stable over time.

## Introduction

Since severe acute respiratory syndrome coronavirus 2 (SARS-CoV-2) started causing clusters of severe respiratory illness in Wuhan, China, in late 2019,^1^ the virus has had a drastic impact worldwide. As of 28 June, there have been 9.8 million confirmed cases reported and almost 496,000 reported deaths.^2^ Unprecedented control measures, such as national lockdowns, have been widely implemented to contain the spread of the virus in a, likely successful,^3-5^ attempt to prevent the collapse of healthcare systems and even larger numbers of deaths among those infected with SARS-CoV-2. Although such measures are important for control of the pandemic, they also affect the economy, unemployment rates, and global supply-chains.^6,7^ While an uncontrolled epidemic has potentially even more drastic consequences,^6^ politicians have to continuously make the difficult decision between continuing strict control measures or relaxing them in some way that would be safe enough in medical terms yet beneficial more broadly across society.

Policy makers and expert groups advising governments about the continued need for social distancing measures in the overall population, specific subgroups and geographic areas heavily rely on accurate and up-to-date information about the prevalence and incidence of people testing positive for COVID-19. In England, a five-level alert system has been introduced, with levels guiding lockdown, social distancing, and relaxation of restrictions decisions.^8^ The levels are largely determined by the effective reproduction number (R) and the number of confirmed cases at any one time.

Here we report the results from the Office of National Statistics (ONS) Coronavirus Infection Survey (CIS) Pilot. This national survey, collecting throat and nose swabs and accompanying data from individuals in households across England, estimates the prevalence and incidence of SARS-CoV-2 in the community in England, and hence directly informs decisions about continued need or relaxation of COVID-19 related control measures.

## Methods

Data were collected between 26 April and 28 June 2020 from individuals that recently took part in the Labour Force Survey or another online survey from the ONS, had consented to be approached for future research, and provided written informed consent to participate in the CIS. At the start of the pilot study, approximately 20,000 households were invited to take part, anticipating approximately 21,000 participants from approximately 10,000 households. After 5 weeks, further households were approached on a weekly basis. As of 28 June, about 45,165 households had been invited to take part, of which approximately 18,894 households were enrolled (not yet all providing data). Parents/carers provided written informed consent for those under 16 years; those aged 10-15 years provided written assent. Only individuals aged 2 years and older living in private households were eligible for inclusion in the survey, meaning that people in care homes, other communal establishments and hospitals are not captured. Because of their selection by ONS for participation in previous studies, households were broadly representative of England. If one or more individuals from a household agreed to participate, a visit by a study worker was arranged and, after signing consent forms, individuals were asked about any symptoms and contacts, together with information about their gender, age, ethnicity and occupation. The study worker provided instructions on how to self-swab the nose and throat, which has been shown to be comparable or even more sensitive than swabs performed by healthcare workers.^9^ Parents/carers took swabs from children under 12 years old. The nose and throat self-swabs were couriered directly to the National Biosample Centre at Milton Keynes, where the samples were tested for the presence of SARS-CoV-2 using reverse transcriptase polymerase chain reaction (RT-PCR) as part of the national testing programme.^10^ After the first visit participants were asked whether they were willing to participate in further follow-up visits: every week for the first 5 weeks of the study, and then monthly for a period of 12 months in total. Therefore, longitudinal follow-up was available for a subsample of included participants.

The project has been reviewed and given ethical approval by South Central - Berkshire B Research Ethics Committee (20/SC/0195).

### Trend in number of positive tests over time

We analysed the proportion of the private-residential population testing positive for COVID-19 from nose and throat swabs over time using dynamic Multilevel Regression and Poststratification (MRP). MRP is an effective method for adjusting a survey sample to be more representative of the population for a set of key (socio)demographic and geographic variables. The method was introduced by Gelman and Little^11^ and has subsequently been further developed, including a dynamic approach that allows assessment of changes over time using all the available data at once.^12^ MRP is increasingly used to make survey samples more representative of (sub)populations and has recently been adopted by the Center of Disease Control (CDC) to provide model-based estimates of the prevalence of diseases at both national level as well as smaller geographical areas in the United States (https://www.cdc.gov/500cities/methodology.htm). In a recent simulation study MRP was found to be superior in terms of accuracy and precision at both the national and regional levels compared to classical survey weighted and unweighted approaches.^13^

MRP generally consists of two steps. First, a multilevel regression model is used to generate the outcome of interest as a function of (socio)demographic and geographic variables. Next, the resulting outcome estimates for each demographic-geographic respondent type are poststratified (weighted) by the percentage of each type in the actual overall population.^11^

We first used a Bayesian multilevel generalised additive regression model to model the swab test result (positive/negative) as a function of age, sex, time and region. Because there were very few missing values (≤ 1%) in these factors, we restricted all analyses to observations with non-missing data. A complementary log-log link was used due to the ability to interpret regression coefficients as arising from an infection process with varying levels of exposure (see Supplementary File 1).^14^ MRP models with random effects for individual participant and/or household nested within region did not converge, meaning that the resulting models could not take into account correlation within individuals or household. However, a model with only one participant sampled from each household gave similar results with somewhat wider 95% credible intervals mainly due to the halved sample size (Supplementary File 1, Fig S1). Time, measured in days since the start of the study (26 April 2020), was modelled using thin plate splines. We set k, the number of basis functions, to five to control the smoothness of the fitted function.^15^ Because variation in trends over time was a key question, we allowed the effect of time to vary by region.

Subsequently, we poststratified the resulting positivity estimates for each demographic-geographic respondent type by the percentage of each type in the overall population and in each region. The post-stratification table was created based on 2011 Census data with age categorised into 5 age-categories (2-11, 12-19, 20-49, 50-69, 70+), and 9 regions within England (North East, North West, Yorkshire and The Humber, East Midlands, West Midlands, East of England, London, South East, South West). This analysis was performed using the rstanarm package in R version 3.6.1.^16^

### Growth rate

To assess to what extent the prevalence was increasing, decreasing or stabilising we estimated the instantaneous growth rates for each region. The growth rate r(t) was estimated using the first and second derivative of the time smoother (s(t), the thin plate spline) from the Bayesian regression with complementary log-log link, following the following formula (derived in Supplementary File 1):

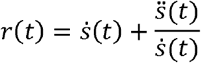

where an over-dot indicates the first derivative and a double over-dot indicates the second derivative. The associated doubling time (or halving time in case of decreasing growth rate) is then:

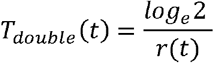

### Associations between variables and testing positive

To assess whether particular subgroups are more likely to test positive for SARS-CoV-2 viral RNA we performed an additional analysis including variables on which we did not post-stratify: work location (working from home, working outside of your home, working from home and working outside of your home, or not applicable), having a job in healthcare or social care that was directly involved with patients/care-home residents, ethnicity (white, Asian, black, mixed, other), household size, and number of children in household (with the latter two variables both obtained from previous underlying ONS surveys). Given the short timescale of the study, and the fact that questions were not always asked at every visit we carried non-missing data forwards and backwards to adjacent visits with missing data. After this, there were very few missing values (≤ 1%), so we again we restricted all analyses to observations with non-missing data only. We used the same model as for the dynamic MRP but in working-aged individuals only (16-74 years inclusive as defined in the Labour Force Survey) with these additional variables included as fixed covariates and age modelled as a continuous variable, using a thin-plate spline (again with k=5), instead of a categorical variable.

Since 8 May 2020 we also collected information on contact with hospitals and care homes in the past 2 weeks. To assess whether having been in the hospital or in a care home was associated with an increased risk of testing positive, we performed another regression also including these two additional variables (participant has had contact; participant has not had contact but someone else in their household has; no one in the household).

In addition, for strong risk factors with ≥ 5 positives in each category we evaluated whether there were different trends over time within each category by including an interaction between time and the respective variables.

### Presence of symptoms among those testing positive

The percentage of participants reporting any symptoms on the day the swab was taken (‘at visit’) or at the visit the week before or after (‘around visit’) was also evaluated for positive and negative tests. Credible intervals for crude proportions were estimated using a Beta(1,1) prior, acknowledging the wide range of estimates of the percentage of infected persons that is asymptomatic in the literature.^17^ To assess the impact of false-positive tests on individuals testing positive without reporting any symptoms, we assessed whether variables that had a clear positive association with testing positive had no or a very small effect on the risk of testing positive while being asymptomatic. This was done by fitting a Bayesian ordinal (adjacent category family) regression model with the following outcomes: (1) negative test; (2) positive test and no reported symptoms; and (3) positive test and reported symptoms. The model allowed for category-specific effects for the variables with clear evidence of an effect on a positive test (regardless of presence of symptoms). This ordinal regression model was fitted using the brms package in R version 3.6.1.^18^ In addition, we evaluated whether the proportion of individuals reporting no symptoms around the visit was stable over time using a Bayesian generalised additive model with a complementary log-log link and time as the only covariate. If false positives were playing an important role in our study, the percentage of individuals not reporting symptoms among those testing positive should increase when the true prevalence is declining.

### Incidence

The daily incidence of new SARS-CoV-2 positive tests among those at risk – i.e. not having tested positive for SARS-CoV-2 before during the study period – was calculated between 12 May and 28 June. We excluded the period before 12 May as for the first 2 weeks of follow-up the first time that an individual could be observed to have become positive was at their first visit (scheduled 7 days after enrolment) and recruitment increased substantially over this period. Given the low number of incident cases a simple Bayesian generalised additive Poisson model with the number of persons at risk on each day as an offset without any covariates other than time was used and no post-stratification was used to adjust for potential unrepresentativeness. The effect of time was modelled using a thin-plate spline (k=5).

### Patient and public involvement

No patients were involved in setting the research question or the outcome measures, nor were they involved in developing plans for design or implementation of the pilot study. No patients were asked to advise on interpretation or writing up of results. Test results are communicated to patients via their General Practitioner. Results are being disseminated to relevant patient communities on an ongoing basis through news media (https://www.ndm.ox.ac.uk/results & https://www.ons.gov.uk/peoplepopulationandcommunity/healthandsocialcare/conditionsanddiseases/bulletins/coronaviruscovid19infectionsurveypilot/previousReleases).

## Results

Figure 1 shows the population representativeness of our underlying survey sample of 34,992 individuals from 16,722 households recruited and providing data from 26 April to 28 June 2020, inclusive in terms of age, sex, and region. Individuals from London and children and younger adults were somewhat underrepresented in our sample, while individuals from all other regions were slightly overrepresented with a more pronounced overrepresentation of those aged 50 years or older. The percentage of males in the survey was very similar to the percentage in the general population of England.

**Fig 1.**
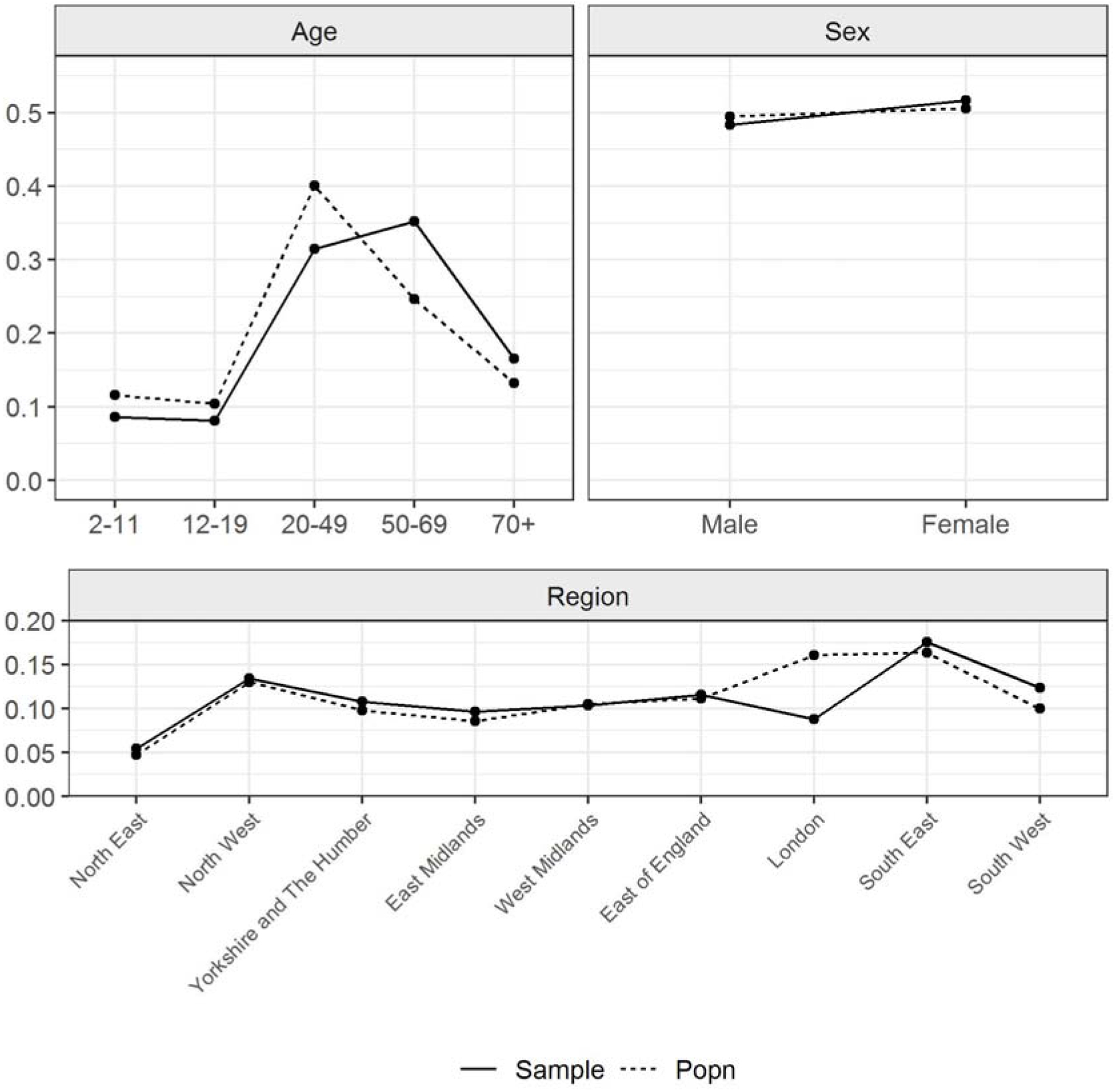
Representativeness of sample in terms of age and sex. Proportion of sample (solid line) and population in England (dashed line) within age- and sex-categories and regions.

Overall 4,443 (13%) individuals contributed only a single test result to the analysis, with the remaining participants contributing a median of 4 (interquartile range 2 to 5). In total results were available from 130,705 tests of nose and throat swabs, of which 134 (0.10%) were positive overall from 112 individuals in 96 households (including repeated positives in some individuals).

After making our results representative of the population by accounting for age, sex, and region using MRP, we observed a decline in the percentage of individuals in England testing positive over time (Figure 2). Specifically, this declined from 0.32% (95% credible interval (CrI) 0.19% to 0.52%) on 26 April 2020 (the first day of the survey) to 0.08 (95% CrI 0.05 to 0.12) on 28 June 2020. Extrapolating to the number of inhabitants in England, this corresponds to 175,000 (95% CrI 105,000 to 284,000) positives on 26 April and 41,000 (95% CrI 26,000 to 64,000) positives on 28 June in England.

**Fig 2.**
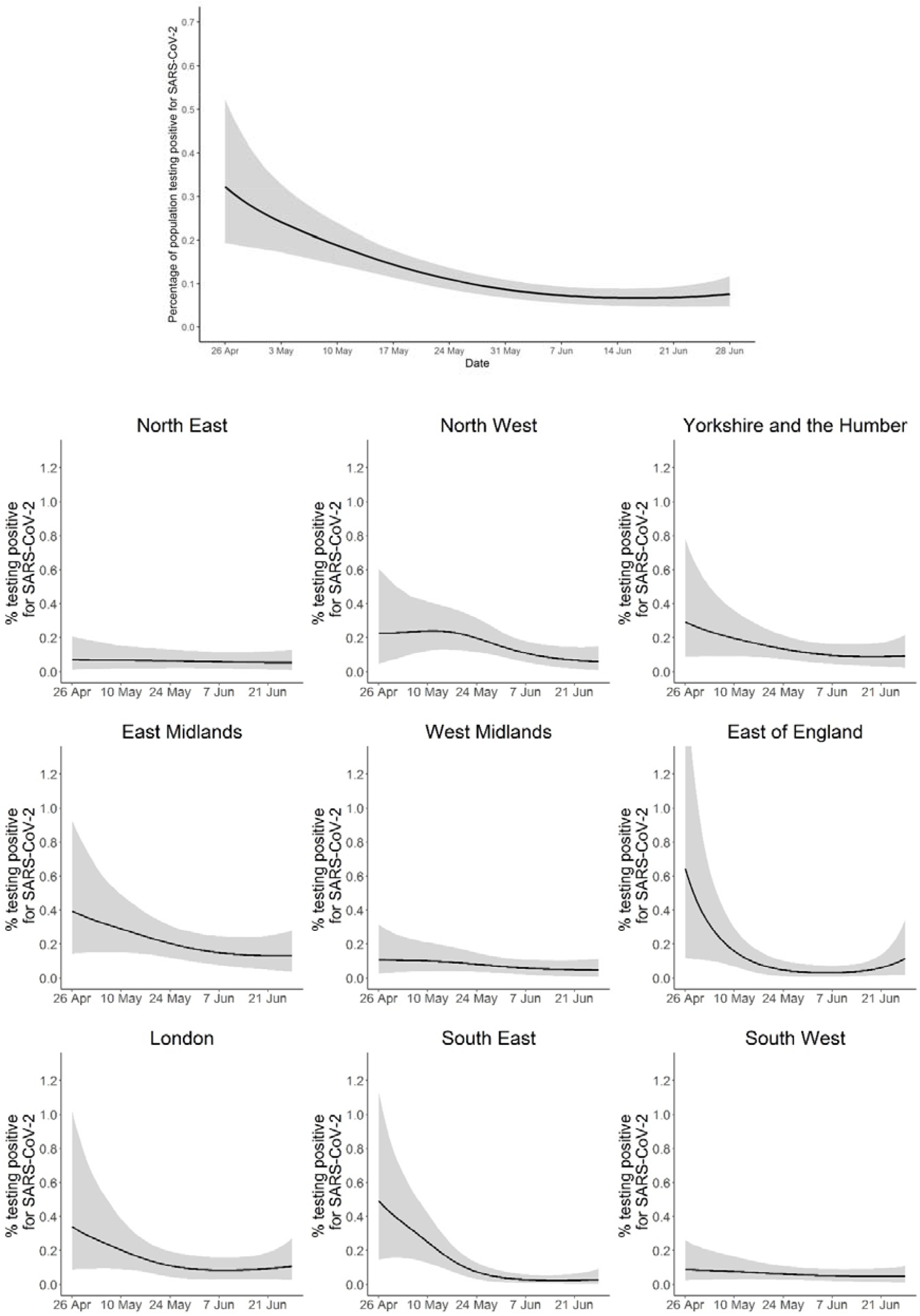
Percentage of population testing positive for SARS-CoV-2 over time. (Top) In England. (Bottom) Within the nine regions of England. Credible intervals are truncated at 1.35% to ensure all regional plots are on the same scale.

The change over time varied somewhat in different regions (Figure 2). For example, the probability that the prevalence in the East Midlands was higher than the average in England on the 28^th^ of June (0.08%) was 88%. However, the credible intervals were wide for most trends within regions, especially where positivity rates seemed to follow a non-linear trend. Figure 3 shows the growth rates over time for England as a whole and the 9 regions. For England as a whole the growth rate was generally negative, consistent with the observed decline in positivity, although it appeared to be levelling off near the end of the study period.

**Fig 3.**
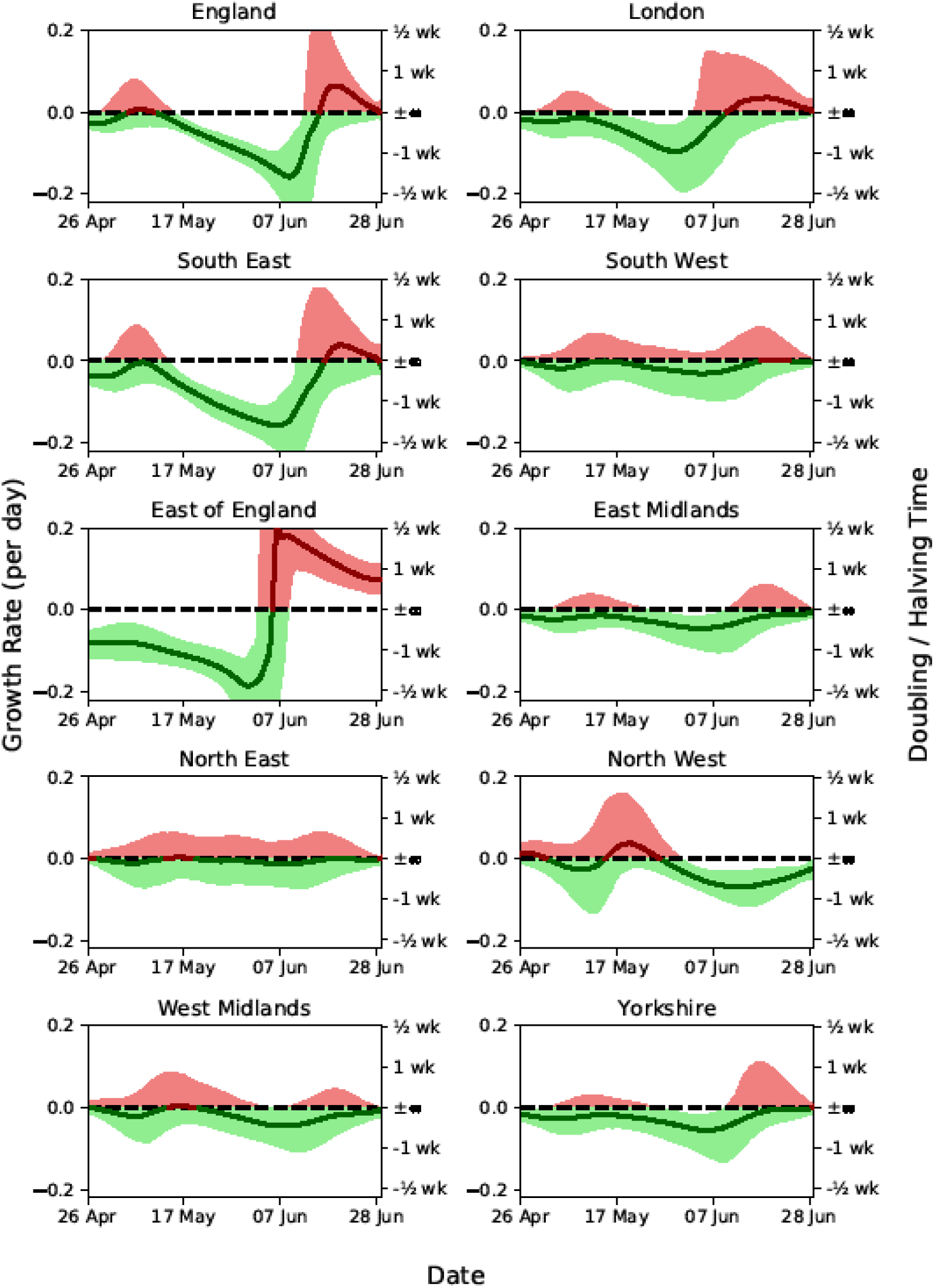
Estimated epidemic growth rates within England and its nine regions, shown as posterior median and 25-75th percentiles. Growth rates are related to doubling (positive weeks) and halving (negative weeks) times.

We also evaluated to what extent different factors, including work-related factors, were associated with increased probability of testing positive for SARS-CoV-2 (Table 1) in individuals aged 16 to 74 years old. There was clear evidence that having a job which involved direct contact with patients and working outside of the home in general was associated with an increased risk of testing positive (Table 1). In addition, individuals that indicated that the work location was not applicable (including not working, retired, students) had a higher risk than those that worked from home. Individuals from ethnicities other than Asian, black, white or mixed also tested positive more frequently although uncertainty was high in this small group which could not be split further. Furthermore, these cases were strongly clustered within households. For the remaining factors there was no clear evidence that there was a positive or negative association with testing positive for the presence of SARS-CoV-2 viral RNA, although individuals that directly work with residents in nursing homes or residential care homes may also have an increased risk (relative exposure (RE) 2.35, 95% CrI 0.85 to 5.27). Within this population of working age, there was no evidence of an effect of age after accounting for other variables (Supplementary File 1, Fig S2). Analyses using an additional random intercept per individual showed similar results, with the same factors identified as being clear predictors of test positivity (Supplementary File 2).

**Table 1.**

| **Factor** | **Number of visits in sample**<br>**(number positive)** | **Relative exposure to SARS-CoV-2**<br>**(95% CrI)<sup>a</sup>** |
| --- | --- | --- |
| Male | 44,308 (53) | Ref. |
| Female | 49,308 (57) | 0.84 (0.57 to 1.25) |
| *Work location* |  |  |
| Working from home | 22,392 (11) | Ref. |
| Working outside of your home | 20,621 (45) | 2.47 (1.40 to 4.55) |
| Both | 4,370 (5) | 1.43 (0.53 to 3.54) |
| Not applicable | 46,233 (49) | 2.09 (1.20 to 3.75) |
| *Job with direct contact with patients or care home residents* |  |  |
| Non-patient facing | 89,643 (87) | Ref. |
| Patient facing | 3,973 (23) | 4.06 (2.37 to 6.72) |
| Non-resident facing | 92,690 (105) | Ref. |
| Care home resident facing | 926 (5) | 2.35 (0.85 to 5.27) |
| *Ethnicity* |  |  |
| White | 88,793 (93) | Ref. |
| Asian | 2,514 (8) | 1.89 (0.87 to 3.64) |
| Black | 788 (2) | 1.04 (0.28 to 3.07) |
| Mixed | 1,068 (0) | 0.46 (0.09 to 1.84) |
| Other | 453 (7) | 7.50 (2.86 to 16.50) |
| *Household size* |  |  |
| 1 | 13,096 (16) | Ref. |
| 2 | 40,426 (28) | 0.62 (0.35 to 1.09) |
| 3 | 17,125 (33) | 1.51 (0.83 to 2.73) |
| 4 | 16,704 (21) | 1.36 (0.68 to 2.63) |
| 5 or more | 6,261 (12) | 1.25 (0.51 to 2.88) |
| *Number of children in household* |  |  |
| 0 | 64,917 (70) | Ref. |
| 1 | 11,206 (22) | 1.01 (0.59 to 1.73) |
| 2 | 10,083 (8) | 0.44 (0.20 to 0.94) |
| 3 or more | 2,593 (8) | 1.65 (0.64 to 4.17) |

<sup>a</sup>A relative exposure of 1 is the reference value (no effect).

An additional analysis on more recent data including information on contact with hospitals or care homes in the previous two weeks showed that having had hospital contact was associated with testing positive for the presence of SARS-CoV-2 viral RNA (RE 2.18, 95% CrI 1.09 to 4.18). Positive tests were also higher if another member of the household had hospital contact but not the person providing the swab themselves (RE 1.99, 95% CrI 0.86 to 4.13). There was no evidence that recent direct or indirect contact with a care home was associated with risk (RE 0.77, 95% CrI 0.18 to 2.79 for having had contact individually and RE 0.48, 95% CrI 0.10 to 1.88 for a household member).

A sensitivity analysis evaluating whether the risk of testing positive declined less in certain risk-groups showed that there was a consistent decline across the board (Supplementary File 1, Fig S3-S4).

### Positive tests with and without reported symptoms

In total, 98 out of 133 (73%, 95% CrI 66% to 81%) visits where individuals had a positive test reported no symptoms on the day the swab was taken, decreasing to 82 positive tests (61%, 95% CrI 53% to 69%) when also including symptoms at visits the week before and after testing positive. Among the subgroup of individuals of working-age (16-74 years old), 66 out of 111 (59%, 95% CrI 50% to 68%) positive tests were obtained from individuals reporting no symptoms around the test. Given the low number of positives that also experienced symptoms, we only included work location, having a job involving direct contact with patients, having a job involving direct contact with care home residents, and region in a multivariable Bayesian ordinal regression model allowing for different effects of factors on the ordinal outcome. Having a job involving direct contact with patients and working outside of the home were both associated with an increased risk of testing positive without reporting symptoms (RE 3.95, 95% CrI 2.13 to 7.06 and 2.43, 95% CrI 1.31 to 4.52 respectively). There was no clear evidence that these associations were stronger for symptomatic than for asymptomatic infections (RE 1.01, 95% CrI 0.47 to 2.06 and RE 1.25, 95% CrI 0.53 to 3.21, respectively). There were insufficient number of positive tests among people having a job involving direct contact with care home residents to draw conclusions about a difference in effect on asymptomatic infection and symptomatic infections (RE 1.33, 95% CrI 0.33 to 4.07 and RE 2.18 95% CrI 0.77 to 5.19, respectively).

The percentage of participants reporting symptoms among individuals testing positive for the presence of SARS-CoV-2 viral RNA remained stable throughout the study period, despite a clear decline in the total number of positives (Supplementary File 1, Figure S5).

### Incidence of positive tests

The incidence of new positive tests among those still at risk appeared to decline initially with a stabilisation or slight increase in new cases near the end of the pilot study (Figure 4). However, 95% credible intervals were wide. Extrapolating to the number of inhabitants in England – ignoring the negligible small fraction of our sample was considered not at risk due to a previous positive test during the survey - this corresponds to 3,400 (95% CrI 1,500 to 6700) new positives per day in England on 28 June 2020.

**Fig 4.**
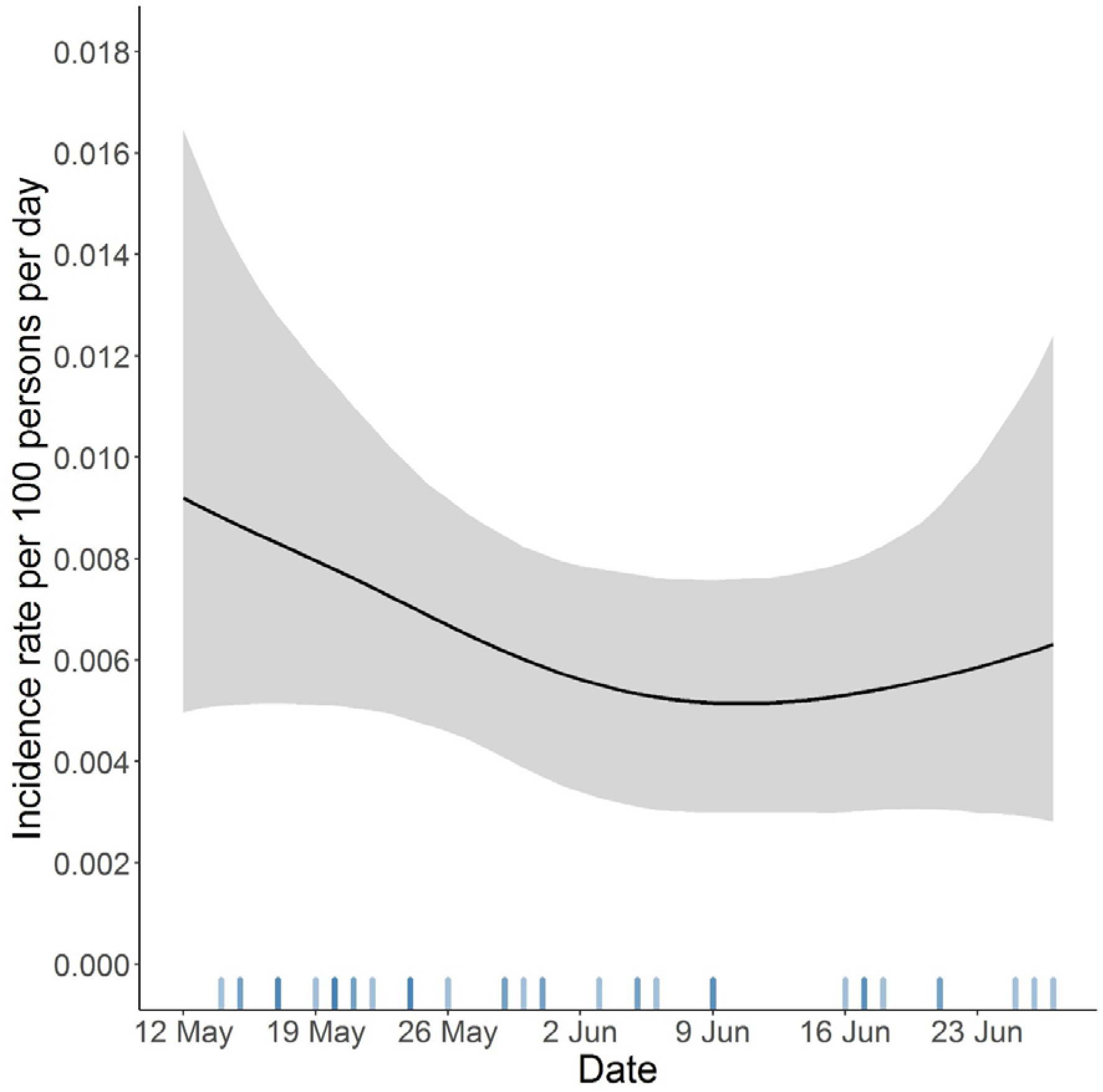
Incidence rate of new SARS-CoV-2 positive tests per 100 persons per day in England. The grey bands represent 95% credible intervals. The intensity of the lines at the bottom of the plot reflects the number of new positive tests recorded each day.

## Discussion

Here we demonstrate a reduction in the percentage of people in private-residential households in the community in England testing positive for SARS-CoV-2 between 26 April and 28 June 2020, suggesting that the lockdown imposed on 23 March 2020 had its intended effect.^19^ Mechanistic dynamic transmission models, calibrated to our nationally and regionally representative estimates of infections, can be used to explicitly estimate the impact of different policy interventions and behaviour changes on the observed and forecasted future trends of the epidemic. As such, our estimates have been regularly updated and shared with the UK Government and Scientific Advisory Group for Emergencies (SAGE) sub-group Scientific Pandemic Influenza sub-group on Modelling (SPI-M) to directly inform decisions about potential changes to the current alert level or relaxation of certain restrictions. However, our survey is not intended to disentangle what interventions or voluntary behaviour changes directly underly any trends.

Notably, we found that a substantial proportion (61%, 95% CrI 53% to 69%) of individuals that tested positive did not report any symptoms on the day of the visit or at visits one week before or after the swab was taken.

Factors associated with an increased risk of testing positive for SARS-CoV-2 viral RNA and reasonable tight 95% credible intervals included working outside the home, having a job with direct patient contact, and having had (indirect) contact with a hospital in the previous two weeks. Because there were relatively few positive cases, we may not have found evidence of an effect for other factors that make a person truly more likely to become infected with SARS-CoV-2. For example, with increasing sample size and/or an increase in the prevalence or using a more informative prior based on external evidence,^20^ it is likely that the evidence of an increased risk among individuals that directly work with residents in nursing homes or residential care homes would have become more apparent (RE 2.35, 95% CrI 0.85 to 5.27 using the current data). Other studies, including the Vivaldi study from the England, have shown that having a resident-facing job likely substantially increases the risk of acquiring an infection when the virus is circulating in a care home.^20,21^

### Comparison with other studies

Our finding of a temporal reduction in the percentage of people in the community infected with COVID-19 in England is consistent with reductions observed in the weekly number of excess deaths in the UK recorded by ONS during the same period.^22^ Similarly, the COVID-19 Hospitalisation in England Surveillance System (CHESS) indicates that during our study period the number of COVID-19 hospital admissions also decreased.^23^ An important advantage of our current study is that it is based on a representative sample of the population, with further correction for potential non-representativeness using MRP. A population-based sample may enable earlier detection of increases and decreases in the epidemic not purely driven by hospital or care home outbreaks than surveillance based on hospitalisations and deaths, which inevitably suffers from a lag between acquiring an infection in the community and potential subsequent hospitalisation and/or death. Furthermore, our national survey is more capable of detecting potential increases in the community among individuals that are less likely to require hospital admission, but potentially still able to contribute to spread of the virus, including if asymptomatic.^24^

There are a few other studies that aimed to assess the prevalence of SARS-CoV-2 infection in the general population. In a study from Vo, an Italian town with a population of 3275 individuals, Lavezzo et al. surveyed 85.9% and 71.5% of the inhabitants at the start and end of lockdown of the town respectively, with an initial infection prevalence of 2.6% (95% confidence interval (CI) 2.1 to 3.3%) and 1.2% (95% CI 0.8 to 1.8%) 14 days later.^25^ The percentage of those who tested positive that did not report any symptoms was 41.0% (95% CI 29.7 to 53.2%) in the first survey and 44.8% (95% CI 26.5 to 64.3%) in the second survey.

As part of a larger study from Iceland, a randomly selected sample of inhabitants aged between 20 and 70 years of age were invited to participate in a survey.^26^ By 4 April, 2283 (33.7%) of those invited had participated, with 13 testing positive (0.6%; 95% CI 0.3% to 1.0% estimated using Wilson score intervals based on numbers reported in the paper). However, it is unclear to what extent the participation rate led to a sample that was not representative of the Icelandic population and no correction for population representativeness was performed. Among a larger sample, including participants recruited via an open invitation, which may bias the sample towards people with symptoms, 57% of individuals testing positive reported having symptoms, although 29% of individuals testing negative also reported having symptoms.

While the studies from Iceland and Vo found that around 40-45% of those with a positive test did not report any symptoms,^25,26^ this percentage is higher in our study, being 59% (50% to 68%) among those 16-74 years old. This may be partly due to differences in respondents, such as no children being included in the study from Iceland, chance (given the wide confidence and credible intervals in all studies), differences in definitions of symptoms, over/under-reporting of symptoms or false positive tests. While false-positives may be a concern with a low prevalence - potentially leading to an overestimation of the percentage of truly infected persons that are asymptomatic - the low number of positive tests in our study overall is also reassuring since it indicates that the specificity of the test is very high. Even in a purely hypothetical situation that the virus is not circulating, a test specificity of 99.9% would be associated with an expected number of positive tests that is approximately equal to what we observed over the entire study period.

Furthermore, the strong associations between expected risk factors, such as directly working with patients, and test positivity in combination with no reported symptoms, together with declining positivity rates over time in those reporting and not reporting symptoms, suggest that there may truly be more asymptomatic cases than reported in some other studies.^17^ A recent meta-analysis of studies focusing on close contacts of confirmed COVID-19 cases suggested that only 15% (95% CI 12% to 18%) of infected individuals are asymptomatic.^17^ However, by informing participants that they were recently in close contact with a confirmed COVID-19 case, individuals may be more likely to think that they have experienced symptoms, resulting in recall bias and overestimating the true prevalence of symptoms among a representative sample of infected persons. Although we may have underestimated the true prevalence of symptoms among SARS-CoV-2 cases in the community, partly due to asking about current symptoms at weekly visits (meaning that very transient symptoms only occurring between visits would have been missed), our study adds to the growing evidence that a substantial proportion of SARS-CoV-2 in the community may be asymptomatic. For example, our estimate aligns with a large study from Italy which recently reported that that 69% (95% CI 67% to 71%) of individuals that test positive do not develop symptoms.^27^

## Limitations of this study

An important limitation of this study is that the number of people in the community that test positive is low, limiting power. From a public health perspective this is positive; however it also means that the multilevel regression model used in our analyses was not able to incorporate likely correlation within households, and that the uncertainty around estimates is relatively large. However, randomly selecting one individual per household gave similar results with somewhat wider credible intervals owing to the halved sample size, suggesting within household clustering is not having a large impact on our results. The data reported here is from the pilot phase of our study, and more data, and hence more positive tests in absolute terms, are expected in future waves of the survey. If a second wave of infections occurs in the coming months, particularly in the absence of an effective vaccine, models including more factors and accounting for correlation between observations from the same household would have greater power to delineate the risk of different subgroups of the population acquiring SARS-CoV-2 infection.

A second limitation is that, in the absence of a true gold standard, we do not know the test sensitivity and specificity, making it difficult to assess what the true prevalence is. From i) the low number of positive tests overall, ii) the finding that risk factors for testing positive regardless of symptoms were (equally) strong for testing positive while reporting no symptoms, and iii) the observation that the percentage of individuals reporting no symptoms among those testing positive remained stable over time despite substantial declines in the overall number of positives, we can conclude that the specificity must be very close to 100% and the number of false positives in our sample is likely low. However, the data cannot inform about the test sensitivity without providing a very informative prior on the true prevalence.^28^ Given the low number of positives, even a low sensitivity of 60% (incorporating both test performance and potentially imperfect self-swabbing) and perfect specificity would mean that the estimated prevalence would still have declined from 0.54% on 26 April to 0.13% on 28 June 2020.

## Conclusions

The percentage of individuals from the community in England testing positive for SARS-CoV-2 clearly declined between 26 April and 28 June 2020, although the decline appears to be levelling off near the end of this period. This adds to the increasing body of empirical evidence and theoretical models that suggest that the lockdown imposed on 23 March 2020 had its intended effect. Important risk factors for testing positive were having a job with direct contact with patients and working outside the home more generally, and a substantial proportion of infections were in individuals not reporting symptoms, indicating that continued monitoring for SARS-CoV-2 in the community will be important for potential early detection of increases in infections due to return to work and other relaxations of control measures. The planned increase in survey sample size, and extensions to other areas of the UK, will likely facilitate exploration of important questions such as what are the trends in prevalence and incidence in smaller geographical areas and what specific types of jobs and other factors make individuals from the community more likely to become infected with SARS-CoV-2.

## Data Availability

Data sharing: De-identified study data are available for access by accredited researchers in the ONS Secure Research Service (SRS) for accredited research purposes under part 5, chapter 5 of the Digital Economy Act 2017. For further information about accreditation, contact or visit the SRS website

## Contributors

The study was designed and planned by SW, JF, JB, JN, IB, ID, PB, KBP, and JVR. KBP, TH, NB, IB, HT, JL and ASW contributed to the statistical analysis. KBP drafted the manuscript and all authors contributed to interpretation of the data and results and revised the manuscript. KBP and ASW are the guarantors of the study. All authors approved the final version of the manuscript and agreed to be accountable for all aspects of the work. The corresponding authors attests that all listed authors meet authorship criteria and that no others meeting the criteria have been omitted.

## Data sharing

Data sharing: De-identified study data are available for access by accredited researchers in the ONS Secure Research Service (SRS) for <u>accredited research purposes</u> under part 5, chapter 5 of the Digital Economy Act 2017. For further information about accreditation, contact or visit the <u>SRS website</u>

## Transparency

The guarantors (KBP and ASW) affirm that the manuscript is an honest, accurate, and transparent account of the study being reported; that no important aspects of the study have been omitted; and that any discrepancies form the study as planned (and, if relevant, registered) have been explained.

## Acknowledgements

**COVID-19 Infection Survey Team**

**Office for National Statistics:** Iain Bell, Ian Diamond, Pete Benton, Joshua Abramsky, Daniel Ayoubkhani, Ian Boreham, Heather Bovill, Nikola Bowers, Dominic Brown, Garnett Compton, James Cooper, Emily Connors, Katie Davies, Claire Dawson, Max Engledew, Onyinye Ezeya, Narcisa Florea, David Foster, Leah Harris, Phillippa Haughton, Nigel Henretty, Tony Hitching, Sarah Henry, Lee Hood, Gareth James, Jaya Jassi,, Joe Jenkins, Katherine Kent, Alex Lambert, Maria Ledgeway, James Lewis, Charles Lound, Victoria Masding, Salah Merad, Tibor Mezzei, David Morrissey, Lisa Moore, Ian O’Sullivan, Ellie Osborn, Vasita Patel, Piotr Pawelek, Tristan Pett, Melissa Randall, Craig Smith, Peter Stokes, Eliza Swinn, Esther Sutherland, Heledd Thomas, Tina Thomas, Maggie Vidler, Alison Wiles, Craig Williams, Nick Woodhill.

**University of Oxford:** Ann Sarah Walker, John Bell, Koen Pouwels

**University of Manchester:** Thomas House

**Public Health England:** John Newton, Julie Robotham, Paul Birrell

**IQVIA:** Helena Jordan, Tim Sheppard, Leontia Gribben, Dan Moody, Leigh Curry.

